# EARLY ACQUISITION AND CARRIAGE OF GENETICALLY DIVERSE MULTI-DRUG RESISTANT GRAM-NEGATIVE BACILLI IN HOSPITALISED SMALL VULNERABLE NEWBORNS IN THE GAMBIA

**DOI:** 10.1101/2022.11.16.22282268

**Authors:** Saikou Y Bah, Mariama A Kujabi, Saffiatou Darboe, Ngange Kebbeh, Bunja FK Kebbeh, Abdoulie Kanteh, Ramatouille Bojang, Joy Elizabeth Lawn, Beate Kampmann, Sesay Abdul Karim, Thushan I de Silva, Brotherton Helen

## Abstract

**Aim:** This detailed genomic study aimed to characterise multi-drug resistant-gram negative bacilli (MDR-GNB) intestinal and skin carriage in small vulnerable newborns and their paired mothers at a low-resource African hospital.

**Methods:** This cross-sectional cohort study was conducted at the only neonatal referral unit in The Gambia with genomic analysis at MRC Unit The Gambia at LSHTM. Neonates <2kg underwent skin and peri-anal carriage swab sampling weekly with paired maternal rectovaginal swabs. Prospective bacteriological culture used MacConkey agar with species identification by API20E and API20NE. All GNB isolates underwent whole genome sequencing on Illumina Miseq platform. Multi-Locus Sequence Typing and SNP-distance analysis were used to identify strain type and infer relatedness.

**Findings:** 135 carriage swabs were obtained from 34 neonates and 21 paired mothers (21 neonate-mother dyads), yielding 137 GNB isolates of which 112 were high quality de novo assemblies. Neonatal MDR-GNB skin or intestinal carriage prevalence was 41% (14/34) at admission with 85% (11/13) new acquisition occurring by 7 days. Multiple MDR and ESBL - GNB species were carried by neonates at different timepoints, most frequently *K. pneumoniae* and *E. coli*, with heterogeneous strain diversity, no evidence of clonality and 111 distinct antibiotic resistance genes, mostly Beta-Lactams (*Bla*-AMPH, *Bla-*PBP, CTX-M-15, *Bla-* TEM-105). 76% (16/21) and 62% (13/21) of mothers had recto-vaginal carriage of at least 1

MDR-GNB and ESBL-GNB respectively, most commonly MDR-*E. coli (*76%, 16/21) and MDR-*K. pneumoniae* (24%, 5/21). Of 21 neonate-mother dyads only one had genetically identical isolates (*E. coli* ST131 and *K. pneumoniae* ST3476).

**Conclusion:** Gambian hospitalised small vulnerable neonates exhibit high MDR and ESBL-GNB carriage prevalence with acquisition between birth and 7 days. The heterogeneous strain diversity and lack of matching isolates between mothers and newborns suggests multiple environmental sources may be important in transmission. Larger genomic studies to confirm these findings in similar resource limited settings is foundational to inform targeted surveillance and infection prevention control policies.

What is known:

- MDR-GNB, especially *Klebsiella pneumoniae* and *Escherichia coli*, are important causes of neonatal invasive infections and mortality in Africa, classified by WHO as pathogens of high priority for research
- Neonatal MDR-GNB carriage is a pre-curser for invasive infection, with preterm, low-birth weight neonates (“Small Vulnerable Newborns”) at greatest risk
- Maternal MDR-GNB carriage is a risk factor for neonatal pathogen acquisition in Europe and other well-resourced settings, but a priority evidence gap exists for transmission pathways for small vulnerable African newborns

What this study adds:

- Hospitalised Gambian small vulnerable neonates have high carriage prevalence of MDR- and ESBL-GNB with acquisition occurring between birth and 7 days
- Heterogeneous diversity of *K. pneumoniae* and *E. coli* strains suggests multiple environmental sources with no evidence of clonal outbreak
- Beta-lactamase genes were most commonly identified with high rates of ESBL- and AMP-C gene production
- Despite high maternal MDR-GNB carriage prevalence there is no genomic evidence indicating widespread transmission from mother to newborn

## Introduction

Neonatal mortality remains unacceptably high in many African and Asian countries, accounting for 47% of deaths in children under 5yrs [1]. Invasive infections are an important contributor to neonatal deaths, with a high burden in Africa and high relative risk of mortality [2,3]. Small vulnerable neonates born either premature (<37 weeks gestation) and/or low birth weight (LBW; <2.5kg) are at greatest risk of infections due to impaired innate and adaptive immunity [4], prolonged hospital stay and invasive procedures [5]. Intestinal carriage of pathogens with translocation across the gut wall is associated with late onset infections and inflammatory disorders [6] and premature infant’s skin integrity is typically impaired, providing an additional route for invasive infection.

An estimated 31% of the 690,000 annual neonatal deaths associated with sepsis are potentially attributable to antimicrobial resistance (AMR) [7]. Within gram negative bacteria (GNB), Enterobacterales are the leading cause of severe bacterial infections in African neonates [2,8,9], with *Klebsiella pneumoniae* and *Escherichia coli* most commonly implicated. Multi-drug resistance (MDR) is seen in up to 82% of invasive neonatal GNB in Africa [10,11], with prevalence increasing [11] and management challenges due to limited diagnostics and therapeutic options [7]. Extended Spectrum Beta Lactamase (ESBL)-producing GNB were listed as pathogens of high priority for research and antibiotic development by the World Health Organisation in 2017 [12], and represent a neonatal public health emergency which is critical to address if global targets to reduce neonatal mortality to ≤12/1000 livebirths are to be met by 2030 [13].

Neonatal MDR-GNB carriage is associated with invasive blood stream infections [14–16], yet detailed understanding of how neonates acquire MDR-GNB within hospitals in resource limited settings (RLS) is limited. LBW [17], prolonged hospital stay and antibiotic use are risk factors [18] for neonatal MDR-GNB acquisition and an association between neonatal ESBL-GNB carriage and premature delivery has been reported [19]. In addition, the intestinal colonisation pattern of hospitalised premature neonates differs from that of healthy, term, breastfed infants but there is a paucity of gestational age specific data from the lowest resource settings and most data originates from HIC settings where the bacterial burden differs. Environmental sources of MDR-GNB on African neonatal units have also been described [20], with contaminated fluids, antibiotic vials, equipment and surfaces implicated and linked to outbreaks [21,22]. Maternal colonization is a well-recognised risk factor for neonatal acquisition and infection with gram-positive bacteria such as Group B Streptococcus [23]. However, the role of maternal colonisation in neonatal MDR-GNB acquisition, especially in Africa has not undergone rigorous scrutiny [24], and the relative contributions of vertical versus horizontal transmission is not known [25]. This is an important gap to address for development of targeted infection prevention control strategies to reduce neonatal MDR-GNB carriage and subsequent invasive infection.

This study aimed to characterise MDR-GNB carriage in small vulnerable newborns at a low-resource African neonatal unit (NNU), with exploration of acquisition in relation to maternal carriage using whole genome sequencing (WGS). Objectives included: 1) Determine species specific MDR-GNB and ESBL-GNB carriage prevalence for neonates and paired mothers; 2) Describe strain-specific *K. pneumoniae* and *E. coli* carriage; 3) Describe antibiotic resistance genes and 4) Explore relatedness of *K. pneumoniae* and *E. coli* isolates within neonate-mother dyads

## Methods

### Study Design

This cross-sectional cohort study was conducted from April–August 2017 as part of a feasibility study to inform the design of a clinical trial investigating the effect of kangaroo mother care (KMC) on survival of small vulnerable newborns [26].

### Study setting

Recruitment took place at Edward Francis Small Teaching Hospital (EFSTH) NNU, the national neonatal referral unit in The Gambia. Approximately 1400 neonates are admitted per year [27] from a mixed in-born (∽6,000 births/year in hospital) and out-born (other health facilities or home) population. Neonatal mortality in The Gambia declined from 49 to 26 per 1000 live births between 1990 and 2018 [1], but is still substantially higher than the SDG 3.2 target of 12 per 1000 live births. 12% of Gambian neonates are born preterm [28], 17% LBW [1] and 28% of neonatal deaths are due to infections [29], with likely underestimation of the contribution of infection to mortality of small vulnerable newborns.

The inpatient case fatality rate at EFSTH ranged from 35% (all admissions) to 48% for neonates <2kg from 2010–2014, with prematurity or LBW accounting for 27% of all admissions [27]. At the time of this study, WHO level 2 special newborn care was provided with oxygen via concentrators, phototherapy, access to blood transfusion and intravenous (IV) fluids. Empirical first line antibiotics were IV ampicillin and gentamicin (age <72h), with ceftriaxone and flucloxacillin used for both community onset and suspected hospital acquired infections (HAI) (age >72h). Ciprofloxacin was second line treatment with carbapenems rarely available. Blood cultures were not routinely available. Admission rates ranged from 80 – 100 neonates/month during the quieter dry season (January–August) to 140 -160 neonates/month during the rainy season (September-December)[27].

### Study population

All neonates <2kg admitted during the study period (April–July 2017) were screened for eligibility with inclusion criteria: weight <2kg and admission age <24h. Neonates were excluded if death occurred before or during screening or in the absence of informed consent. Specific exclusion criteria for skin swabs included topical antibiotics or steroids applied to skin since birth, and generalized or local skin disorder within 4cm of swabbing site [30]. Exclusion criteria for peri-anal swabs included imperforate anus or anal stenosis, previous gastrointestinal surgery or diarrhoea within preceding 24h [30]. Mothers were approached for consent to provide rectovaginal swabs once they were available, with exclusion if known HIV infection, major gastrointestinal surgery within previous 5 years, diarrhoea or constipation within preceding 24h, or current sexually transmitted infection [30]. Participant identification was pseudoanonymised using unique identification numbers, with identity known only by the researchers.

### Data collection & procedures

Neonatal swabs, anthropometric and clinical data, gestational age assessment (New Ballard score [31]), antibiotic use and in-hospital outcomes were collected via direct observation, parental interviews and medical record review as soon as possible after admission then weekly (day 7, 14, 21, 28) until discharge or death. Data were recorded electronically using REDCap™. Neonatal skin swabs were obtained by composite sampling from the xiphisternum and peri-umbilical area. Peri-anal samples were taken in lieu of stool or rectal samples for intestinal carriage [32] due to less invasive sampling. Rectovaginal (RV) swabs were taken from mothers. All samples were taken by trained personnel with FLOQ® swabs and stored in Amies transport medium at +4 to +8°C prior to transfer to the Medical Research Council, The Gambia at London School of Hygiene & Tropical Medicine (MRCG) within 48 hrs.

### Microbiological processing

Prospective bacteriological processing and storage of isolates occurred at MRCG, ISO-15189 accredited Clinical Laboratories. Samples were processed immediately by culture on MacConkey agar plates incubated aerobically at 35-37°C, with identification of Enterobacterales and other GNB using API 20E and API 20NE respectively.

### Molecular processing and bioinformatic analysis

Whole genome sequencing was performed at the MRCG Genomics Facility. Deoxyribonucleic acid (DNA) was extracted from cultures of all identified GNB, expanded from single bacterial colonies using the QIAamp DNA extraction kit (Qiagen) following manufacturer’s instructions and Illumina Miseq sequenced to 250 cycles. Quality control and trimming of raw sequence reads were done using FastQC (v0.11.8) and trimmomatic (v0.38) respectively to remove low quality bases and sequencing adapters [33]. Whole genome *de novo* assemblies were generated using SPAdes (kmers: 21,33,55 and 77) and quality checked using Quast [34,35]. All draft assemblies greater than 500 contigs were removed from downstream analyses. Multi locus sequence typing (MLST) was done using the get_sequence_type from the mlst_check from the Sanger institute pathogen group https://github.com/sanger-pathogens/mlst_check). Genomes were annotated using Prokka and core genomes analysed using Roary [36,37]. SNP-dist was used to calculate genetics distances (single nucleotide polymorphisms [SNPs]) between isolates of the same species to infer relatedness. Abricate was used to determine AMR gene carriage using the ARG-ANNOT database setting a minimum coverage of 70% and identity of 75% (https://github.com/tseemann/abricat). Genotypic MDR was defined as presence of 3 or more AMR encoding genes, as classed using MEGARes database [38]. Isolates were defined as ESBL producing if a recognised ESBL gene type was identified on whole genome sequencing. Maximum likelihood phylogenetic trees were generated from aligned core SNPs using RAxML with 100 bootstraps, visualized and annotated in iTOL [39,40].

### Ethics & consent

Ethical approval was obtained from LSHTM Observational Ethics Committee (Ref. 11887) and Gambian Government/MRCG Joint Ethics Committee (Ref. 1503). Written informed consent was sought from the neonates’ first available relative prior to data collection with separate consent for maternal sampling sought from the mother. Consent was sought for future research on samples with exclusion from genomic analysis if not provided. Participants were free to withdraw from the study at any time [26].

## Results

### Enrolment & study participants

Of 89 neonates screened, 36 met eligibility criteria and 34 underwent sampling with 114 neonatal carriage swabs obtained (Figure 1). Twenty-one mothers were sampled, of which 19 were linked to sampled neonates. 21 neonate-mother dyads were included due to presence of two twin-mother pairs. 76% of maternal recto-vaginal samples were taken within 24h of neonatal admission (Table 1).

**Figure 1.**
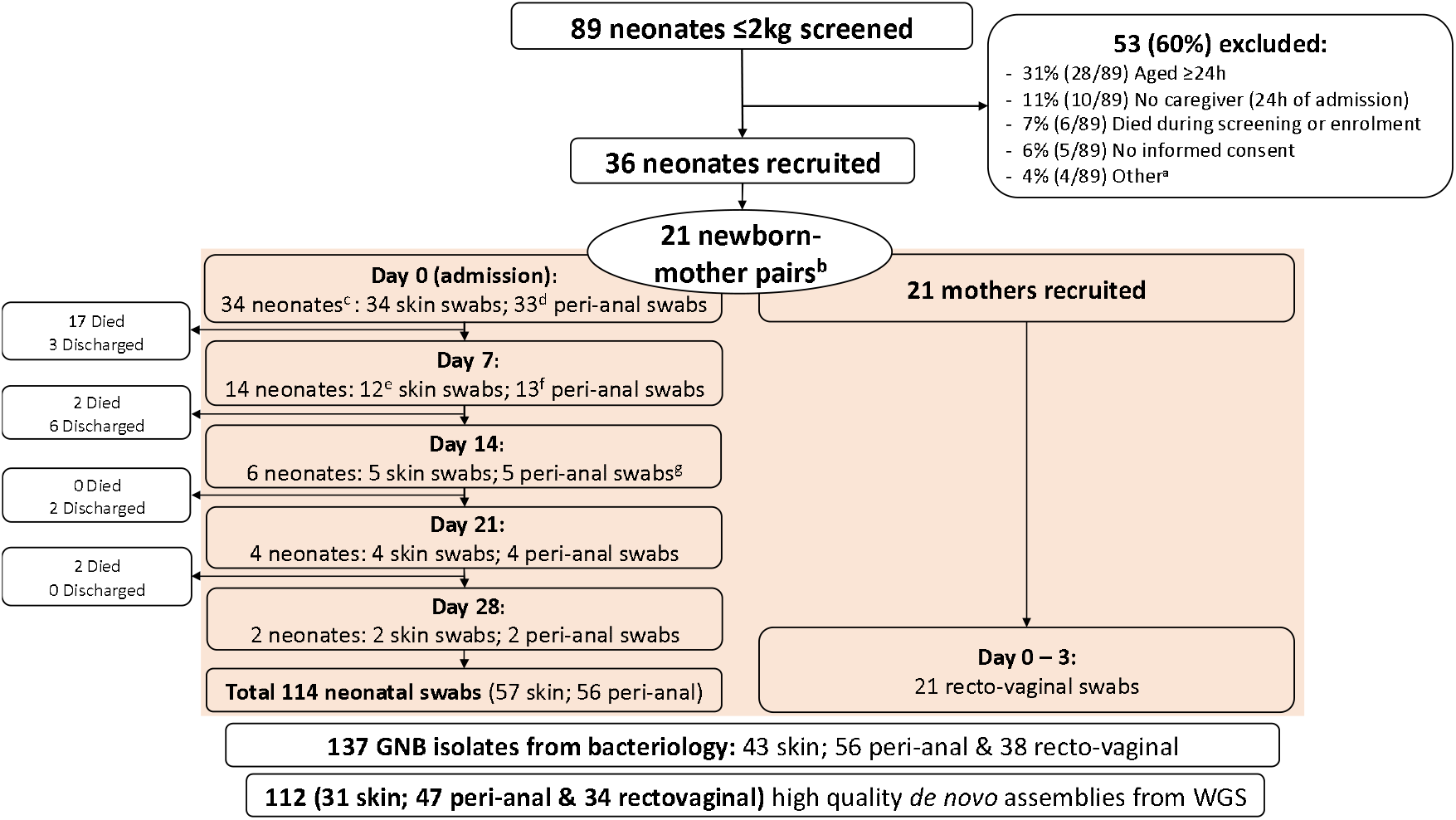
Overview of screening, enrolment, sampling and isolate sequencing for neonatal and maternal participants a) Other reasons for non-recruitment included weight >2kg on study scales (n=2) and no staff available to perform study procedures (n=2) b) Two mothers were sampled in absence of neonatal paired swabs and two sets of twins were included c) 36 neonates were enrolled but 2 were not sampled, due to rapid deterioration and death (n=1) and lack of consent for neonatal sampling (n=1) d) One participant did not meet eligibility for peri-anal samples due to imperforate anus e)Two neonates did not have skin swabs taken: One met exclusion criteria; One had consent withdrawn for repeat sampling f) One neonate did not have peri-anal swabs taken as consent was withdrawn for repeat sampling g) One newborn did not have skin or peri-anal samples taken due to error GNB = Gram-negative bacilli; WGS = whole genome sequencing

The median neonatal admission weight was 1330g (71% <1.5kg) and median gestational age 33 weeks, with 18% (6/34) twin pairs. 91% (32/34) were born in a health facility with a combination of neonates born at the study site (inborn) and elsewhere (out-born) with postnatal transfer. At least one septic risk factor (maternal fever, suspected chorioamnionitis or prolonged rupture of membranes >18h) was present in 14% (3/21) of mothers with 19% (4/21) receiving antibiotics 48h before delivery. The inpatient case fatality rate within 28 postnatal days was 62% (21/34), with median age at death 2.5 days. Reliable data on the cause of death was not available (Table 1).

### Sampling and overview of Gram-Negative Bacilli isolates

135 neonatal and maternal carriage swabs yielded 137 GNB isolates from conventional bacteriology with 112 high quality *de novo* assemblies obtained (Figure 1).

### Spectrum of Gram Negative Bacilli carriage and rates of genotypic Multi-Drug Resistance

Of 112 high quality *de novo* assemblies obtained, 70% (78/112) originated from neonates and 30 % (34/112) from mothers. *E. coli* (40.2%, 45/112) and *K. pneumoniae* (33.0%, 37/112) were the most frequently identified species. Nearly three-quarters of isolates (73%, 82/112) exhibited genetic MDR (*K. pneumoniae* 76% (28/37) and *E. coli* 76% (34/46)). Half (23/45) of the *E. coli* isolates were derived from maternal rectovaginal samples. 76% (28/37) of the *K. pneumoniae* identified were neonatal in origin, with an equal contribution from skin and peri-anal swabs. 82% (23/28) of neonatal *K. pneumoniae* isolates were MDR, predominantly those identified from peri-anal isolates (81%, 17/21). 27% (3/11) of *Acinetobacter baumannii*, 100% (3/3) of *Citrobacter freundii* and 86% (6/7) of *Enterobacter cloacae* were MDR.

### Neonatal & maternal MDR-Gram Negative Bacilli carriage during NNU admission

41% (14/34) of neonates carried >1 MDR-GNB and 38% (13/34) >1 ESBL-producing GNB at time of NNU admission. This was split equally between skin (26%) and peri-anal (24%) (Table 2). All surviving neonates carried >1 MDR-GNB after one week, with 85% (11/13) colonised with at least one new MDR-GNB strain compared to admission (Table 2).

**Table 2.** Genotypic MDR-GNB and ESBL-GNB carriage amongst sampled neonates and mothers from admission to day 7 of hospital admission

|  | Neonates |  |  |  |  |  | Maternal |
| --- | --- | --- | --- | --- | --- | --- | --- |
|  | Total<sup>a</sup> |  | Peri-anal |  | Skin |  | Recto-vaginal |
|  | D0<br>N=34 | D7<br>N=13 | D0<br>n=33 | D7<br>n=13 | D0<br>n=34 | D7<br>n=12<sup>b</sup> | D0 – D3<br>n=21 |
| $\geq 1$ GNB isolated, N° (%) | 19<sup>c</sup> (56) | 13<sup>d</sup> (100) | 12 (36) | 12 (92) | 15 (44) | 5 (42) | 17 (81) |
| $\geq 1$ MDR-GNB, N° (%) | 14<sup>c</sup> (41) | 13<sup>d,e</sup> (100) | 8 (24) | 12 (92) | 9 (26) | 4 (33) | 16 (76) |
| $\geq 1$ ESBL-GNB, N° (%) | 13<sup>c</sup> (38) | 12<sup>d,f</sup> (92) | 6 (18) | 11 (85) | 8 (24) | 4 (33) | 13 (62) |
| *Escherichia coli*, N° (%) | 10 (29) | 4 (31) | 4 (12) | 3 (23) | 7 (21) | 1 (8) | 17 (81) |
| MDR-*E. coli*, N° (%) | 6 (18) | 4 (31) | 3 (9) | 3 (23) | 3 (9) | 1 (8) | 16 (76) |
| ESBL-*E. coli*, N° (%) | 5 (15) | 4 (31) | 2 (6) | 3 (23) | 3 (9) | 1 (8) | 10 (48) |
| *Klebsiella pneumoniae*, N° (%) | 7 (21) | 8 (62) | 4 (12) | 7 (54) | 3 (9) | 2 (17) | 8 (38) |
| MDR-*K. pneumoniae*, N° (%) | 3 (9) | 7 (54)<sup>d</sup> | 1 (3) | 6 (46) | 2 (6) | 2 (17) | 5 (24) |
| ESBL-*K. pneumoniae*, N° (%) | 6 (18) | 8 (62)<sup>f</sup> | 3 (9) | 7 (54) | 3 (9) | 2 (17) | 6 (29) |
| *Acinetobacter* spp., N° (%) | 5 (15) | 2 (15) | 4 (12) | 2 (15) | 4 (12) | 1 (8) | 0 |
| MDR-*Acinetobacter* spp. N° (%) | 1 (3) | 2 (15) | 1 (3) | 2 (15) | 0 | 0 | 0 |
| ESBL-*Acinetobacter* spp. N° (%) | 0 | 0 | 0 | 0 | 0 | 0 | 0 |
| **Other GNB<sup>s</sup> N° (%)** | 8 (24) | 3 (23) | 3 (9) | 3 (23) | 6 (18) | 0 | 2 (10) |
| **MDR-Other, N° (%)** | 7 (21) | 3 (23) | 3 (9) | 3 (23) | 5 (15) | 0 | 2 (10) |
| **ESBL-Other, N° (%)** | 4 (12) | 2 (15) | 1 (3) | 2 (15) | 3 (9) | 0 | 0 |

ESBL = Extended Spectrum Beta Lactamase; GNB = Gram-Negative Bacilli; MDR = Multi-drug resistant;
a. Neonates with either peri-anal or skin sample b. 1 neonate underwent peri-anal sampling at day 7 but met clinical exclusion criteria for skin sampling c. 8 neonates had a GNB identified from both peri-anal and skin samples at first sampling, 3 with a MDR-GNB at both sites and 1 with ESBL-GNB at both sampling sites d. 4 neonates had a GNB identified from both peri-anal and skin samples at day 7, with 3 neonates carrying MDR and ESBL pathogens at both sampling sites e. 85% (11/13) of surviving neonates who were sampled acquired a new MDR-GNB by day 7 of admission with 12 isolates identified (*E. coli* n=2; *K. pneumoniae* n=6; *A. baumannii* n=2; *E. cloacae* n=1; *Pseudomonas aeruginosa* n=1) f. 69% (9/13) of surviving neonates who were sampled acquired a new ESBL-GNB by d7 of admission, with 10 ESBL producing isolates identified (*E. coli* n=2; *K. pneumoniae* n= 6; *E. cloacae* n=1; *Pseudomonas aeruginosa* n=1) g. Other GNB: *Enterobacter cloacae* (n=5); *Citrobacter freundii* (n=2); *Cronobacter sakazakii* (n=3); *Pseudomonas aeruginosa* (n=2); *Pseudomonas putida* (n=1); *Salmonella enterica* (n=1)

29% (10/34) of neonates were colonised with *E. coli* at admission, predominantly on the skin (21%, 7/34), with MDR and ESBL *E. coli* carriage present in 18% (6/34) and 15% (5/34) of neonates respectively. Comparatively less neonates carried *K. pneumoniae* at admission (21%, 7/34), with only three (9%) colonised with MDR *K. pneumoniae*, although 18% (6/34) carried ESBL-producing *K pneumoniae*. The carriage prevalence increased for both MDR and ESBL *E. coli* and *K. pneumoniae* during the first week of admission, with the greatest increase observed for *K. pneumoniae* (MDR from 9% to 54%; ESBL from 18% to 62%) and to a lesser extent *E. coli* (MDR from 18% to 31%; ESBL from 15% to 31%) (Table 2). Eleven neonates had 21 isolates identified from samples taken after day 7 of admission, mostly *K. pneumoniae* (57%, 12/21 isolates), identified predominantly from peri-anal swabs (83%, 10/12 neonates).

76% (16/21) of mothers carried >1 MDR-GNB in their recto-vaginum and 62% (13/21) had an ESBL-producing pathogen. *E. coli* was most commonly identified, with 76% (16/21) of mothers colonised with an MDR-*E. coli*. One quarter (24%, 5/21) of mothers had recto-vaginal carriage of MDR-*K. pneumoniae*.

### Genetic diversity of Gram-Negative Bacilli carriage isolates

#### Klebsiella pneumoniae

We obtained 37 quality *de novo K. pneumoniae* assemblies. Eighteen different sequence types (STs) were determined. ST607 was the predominant ST (11%, 4/37), followed by ST37, ST133 and ST307 (8%, 3/37 each), with 19% (7/37) of isolates not assigned a ST. Seven neonates had multiple *K. pneumoniae* isolated at different time points, of whom four had distinct STs: N019 (3 isolates), N020 (4 isolates), N029 (3 isolates) and N040 (2 isolates). Only two neonates carried a genetically identical *K. pneumoniae* at two or more different time points: 1) A female neonate had *K. pneumoniae* ST502 peri-anal carriage at 20d and 28d (N002; SNP difference=11); 2) A male neonate had peri-anal carriage of *K. pneumoniae* ST607 at 7d and 14d (N012; SNP distance=0). One set of twins carried the same *K. pneumoniae* (ST37, SNP distance=0) on day 8 (N019, twin 1, peri-anal) and day 21 (N020, twin 2, skin). This twin pair also had peri-anal carriage of an identical *K. pneumoniae* (ST476;SNP distance=17) on day 14 (N019) and day 29 (N020), which was not identified in their mother (M009). Furthermore, there was only a single instance of a neonate-mother dyad with identical *K pneumoniae* (ST3476; SNP distance=0), isolated from a neonatal d0 peri-anal swab (N048) and maternal rectovaginal swab (M022)(Figure 2A). This involved a female preterm singleton, admitted following vaginal delivery at another health centre. This newborn died within 7 postnatal days, hence further samples weren’t available.

**Figure 2.**
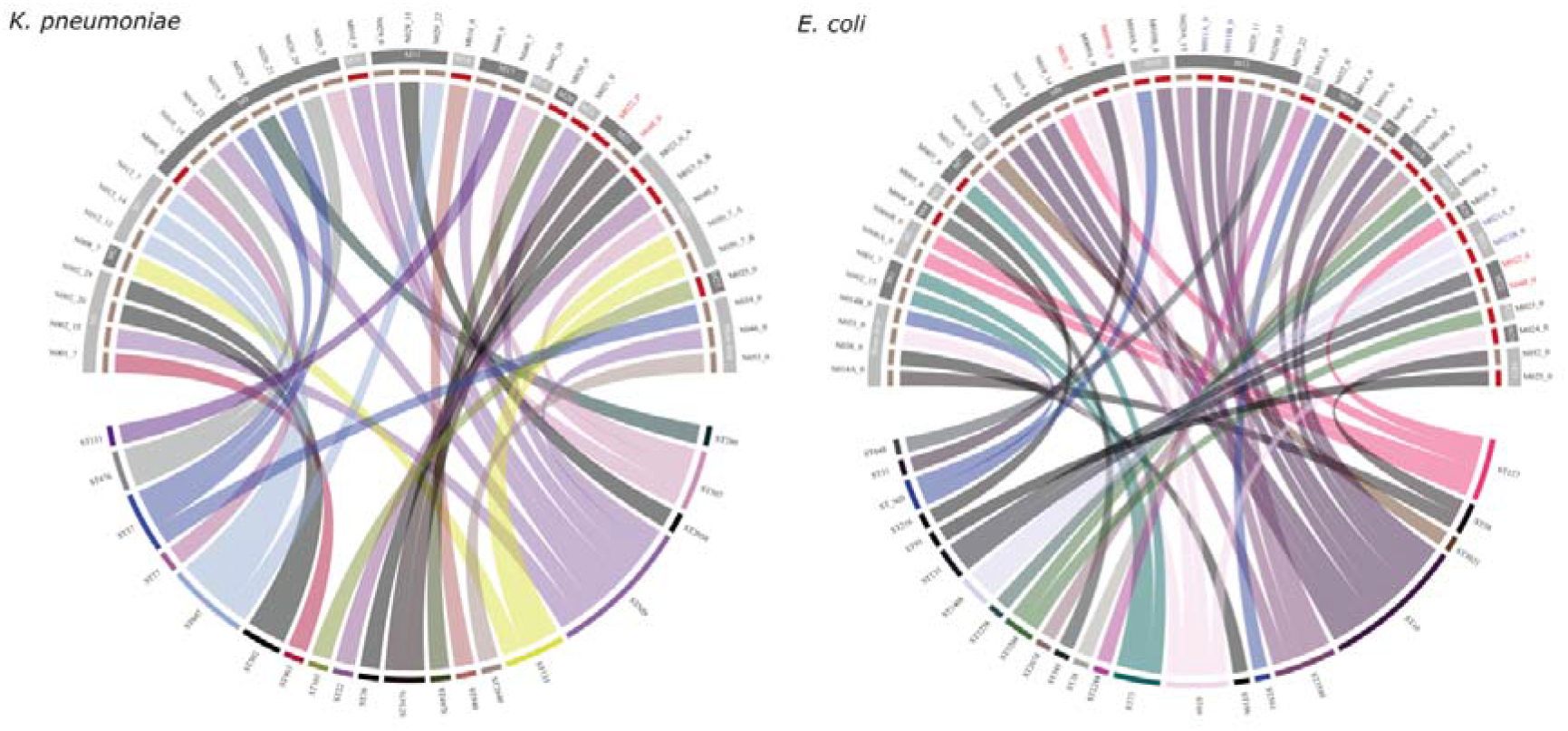
Multilocus sequencing types and genetic relatedness of *K. pneumoniae* (A) and *E. coli* (B). SNP-distance was used to determine genetic relatedness of isolates using the core-genome alignment obtained from Roary. Sequence type (ST) were determine using the mlst package https://github.com/tseemann/mlst. SNP distance were imported into R to generate the circus plots using the circlize package. In the top half of the circus plot, the inner segments indicate whether the isolates were collected from a neonate (brown) or mother (red), mothers’ study ID (MXX) and day of sampling labelled on the outside separated by underscore (_). Sample IDs highlighted with the red font are mother neonate pairs that have the ST. The bottom half indicates the ST types of *K. pneumoniae* or *E. coli*. Connecting lines joining the upper and lower halves of the circus plot indicates to which ST a particular an isolate belongs to.

#### Escherichia coli

45 high quality *E. coli* genomes were obtained with 21 different STs identified and two isolates (4.4%) not assigned a ST. ST10 was most common (20%, 9/45), followed by ST69, ST127 and ST3580 (8.8%, 4/45 each). The most common neonatal derived *E. coli* strains were ST10 and ST3580 (18%, 4/22, each). Three neonates carried multiple *E. coli* isolates at varying time-points: 1) A female singleton weighing 1500g had *E coli* on both skin and peri-anal swabs at d0 (NST58 and unassigned ST; N014); 2) A female twin carried *E. coli* ST10 on day 0 (PA) and day 7 (skin and PA)(SNP distances=1–128; N019) and *E. coli* ST127 was present on day 14 (PA); 3) A male singleton had four identical isolates of *E*. coli ST3580 (SNP distance=0-3; N029) on skin and peri-anal samples taken between day 7 and day 21, with *E. coli* ST648 present on day 21 (Figure 2). *E. coli* ST10 was the most commonly observed isolate in maternal rectovaginal samples (24%, 5/21). 14% (3/21) of mothers carried >1 *E. coli* ST. Only one mother-neonate pair had identical *E. coli* carriage (ST131; SNP distance=0), with neonatal peri-anal carriage at d0 (N048) and maternal recto-vaginal sample (N022) which was obtained within 24h of admission. This mother-neonate pair also had identical *K. pneumoniae* strains, as described above.

### Antimicrobial resistance (AMR) gene carriage

A total of 1,141 AMR genes were identified from 112 isolates, representing 111 distinct gene types, and encoding resistance for 10 antibiotic classes. Beta-lactam resistance was the most common (43.2%, 48/111), followed by resistance to Aminoglycosides (18.2%, 20/11) (Supplementary Table 1 and Figure 2). All *K. pneumoniae* and *E. coli* isolates had > 2 AMR genes, with a median of 11 AMR genes per isolate for both bacteria (*K. pneumoniae* range [3 – 19]; *E. coli* range [5 – 15]).

*K. pneumoniae* isolates had a total of 446 AMR genes (51 distinct gene types), encoding resistance to 10 antibiotic classes, most commonly beta-lactamases (35.9%, 160/446). 19 distinct beta-lactamase genes were identified, most commonly *Bla*AmpH and *Bla*PBP (97% (36/37) and 94% (35/37) of isolates respectively). At least one ESBL genes was present in 92% (34/37) of *K. pneumoniae* isolates, most commonly CTX-M-15 (20/37, 54%) and Bla-TEM-105 (18/37, 49%) (Figure 3, Supplementary Table 1).

**Figure 3.**
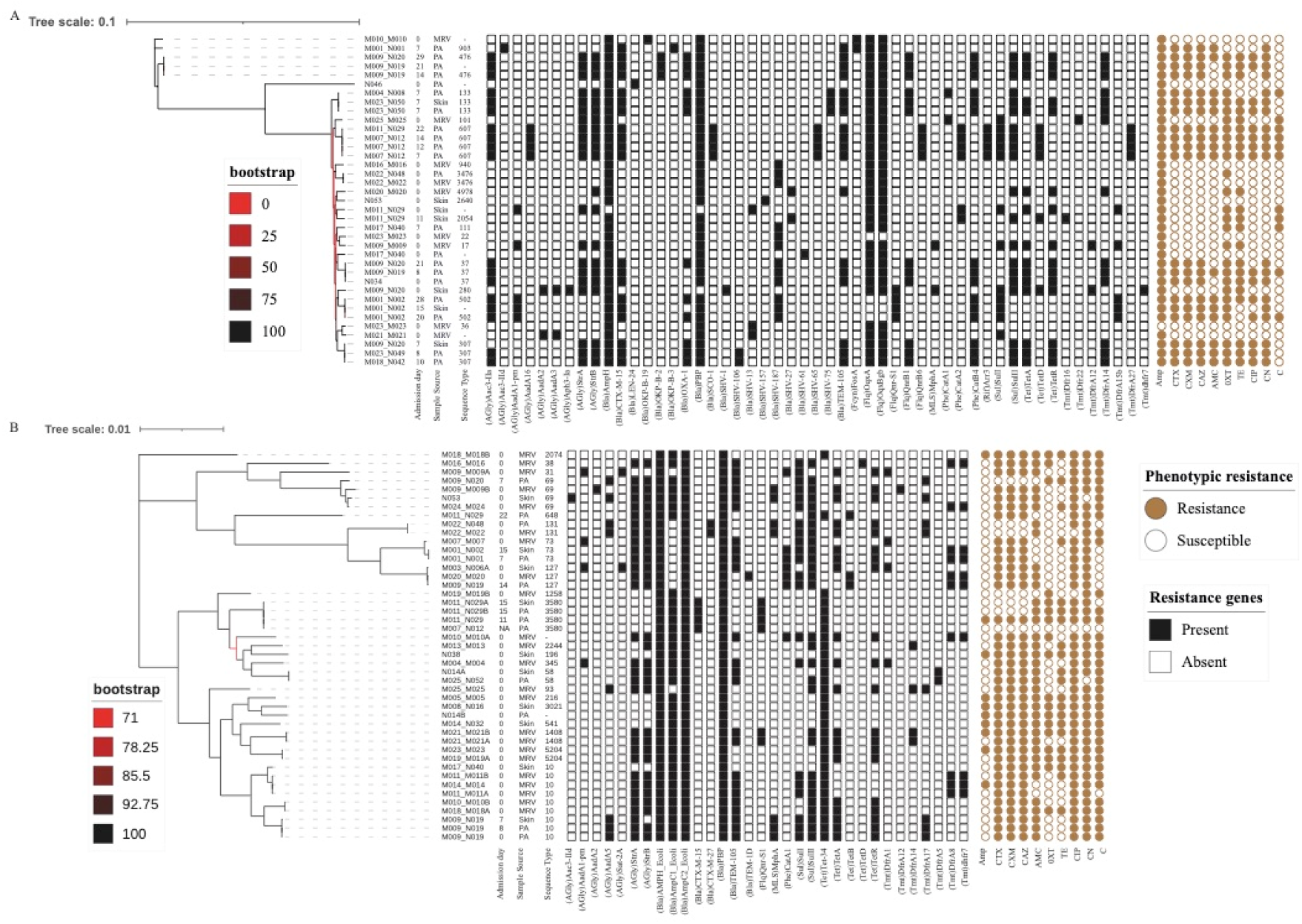
Phylogeny of *K. pneumoniae* (A) and *E. coli* (B) isolates. A maximum likelihood phylogenetic tree was constructed from core-genome SNPs using RAxML with 100 bootstraps. For both species isolates clustered by STs. Also shown are the presence (black-filled square) or absence (white empty-square) of antimicrobial resistance genes (AMR). AMR genes were determined using Abricate using the Argannot database. Phenotypic resistance is shown by closed brown circles and susceptibility denoted by the open brown circles.

*E. coli* isolates harboured 456 AMR genes (32 distinct gene types), encoding resistance to 8 different antibiotic classes, most commonly beta lactamases (44.1%, 201/456). Eight types of beta-lactamase genes were present, with *Bla*AmpH, *Bla*PBP and *Bla*AmpC2 carried by all isolates along with *Bla*AmpC1 in 78% (35/45). Two-thirds (66.7%, 30/45) of *E. coli* isolates harboured an ESBL-gene, mostly Bla-TEM-105 (53%, 24/45) (Figure 3, Supplementary Table 1).

None of the major carbapenemase resistance genes (VIM-, IMP-and NDM-type metallo-beta-lactamases, KPC-nor OXA-48) were identified in *E. coli* and *K. pneumoniae*. However, 91% (10/11) of *A. baumannii* isolates harboured *bla*Mbl, a class B3 beta-lactamase, which has carbapenemase activity (Figure 3, Supplementary Table 1).

## Discussion

We identified high carriage prevalence of MDR-and ESBL-producing GNB in small vulnerable neonates within 24h of NNU admission and extensive acquisition after 7d of hospital stay. Multiple MDR and ESBL-GNB species were identified in individual neonates at different time points, most commonly *K. pneumoniae* and *E. coli* with heterogeneous diversity of strains, no evidence of clonality and a wide range of AMR genes, most commonly Beta-Lactamase genes. Maternal carriage prevalence of MDR-GNB, predominantly *E. coli* was very high, but only one newborn-mother dyad had evidence of genetically identical strains for both *K. pneumoniae* and *E. coli*.

Our observed high rate of in-patient neonatal MDR-GNB acquisition is greater than in European outbreak situations; 24% in Norway [41], middle-income NNUs such as Morocco (58%) [21] and Malaysia (22% and 52%) [42]. Our findings are comparable to phenotypic data from Ethiopia (74% prevalence of ESBL-GNB after 48h of admission)[43] and other West African countries such as Ghana (65%) [44], although the proportion of ESBL-GNB varies considerably within African regions and hospitals, as shown by large heterogeneity in pooled prevalence from East Africa (12 – 89%) [45]. Our cohort of mixed inborn and out-born neonates had high MDR-GNB carriage prevalence at time of NNU admission (44%), suggesting that rapid colonisation occurs during the pre-admission period, although we are unable to comment on precise timing and source of acquisition. Our observed 54% MDR *K. pneumoniae* carriage prevalence after 7 days of admission (all harbouring an ESBL gene) is similar to that reported from a tertiary NNU in Ghana in which 49.6% of neonates had phenotypic MDR *K. pneumoniae* and 75.6% exhibited ESBL activity at median 3d of admission [18]. The high carriage rates at both admission and after 7 days likely reflects varying health system factors such as availability of WASH resources and optimal hand hygiene, provision of sterilisation techniques, over-crowding and under-staffing [46], sub-optimal infection prevention control systems and re-use of disposable consumables and equipment. All these factors have been linked to MDR-GNB outbreaks in other African NNUs [20] and were also observed at this site [22].

MLST identified substantial intraspecies diversity with 21 different *E. coli* sequence types (most frequently ST10, ST69, ST127 and ST3580), and 18 *K. pneumoniae* strains (most commonly ST607, ST37, ST133, ST307). *E. coli* strains range from harmless commensals to pathogenic variants associated with invasive infections [47] and several strains colonising our cohort have been linked to neonatal infections in Africa and Asia (ST10, ST69) [47]. We identified *E*.coli ST131 in 4% (2/45) of *E*.*coli* isolates (one each in a neonate-mother dyad), important as it is a highly virulent strain and major neonatal pathogenic variant [47]. This finding is consistent with a community study in Guinea Bissau which identified ST131 in only 4% of *E*.*coli* isolates from >400 children, including neonates [48]. Two of our most frequently identified *K. pneumoniae* strains (ST37, ST307) are associated with invasive neonatal infection and have been previously reported from Ethiopia (ST37), Rwanda (ST307) and Nigeria (ST307) [47]. We did not identify any *K. pneumoniae* ST39 or ST31535 (*K quasipneumoniae)*, which were both implicated in contemporaneous outbreaks at the site one year previous to our sampling period [22], suggesting that prior outbreaks were contained. There are limited other African genomic data describing strain specific neonatal *E. coli* and *K. pneumoniae* carriage and this is a priority research area.

Beta-lactamases were the most common AMR genes identified, with a predominance of AmpH, penicillin binding protein (PBP), TEM and CTX-M gene types, conferring resistance to ampicillin and 3^rd^ generation cephalosporins (first and second line antibiotics) in 43% of gram-negative isolates. The low prevalence of carbapenem resistance genes in our isolates contrasts with higher levels reported from Ghana (15.6%) [49], and Thailand (64%) [50] likely reflecting the limited availability of carbapenem antibiotics in our setting and thus reduced selective pressure. However, nearly ubiquitous presence of *blaMbl* in *A. baumannii* is of concern due to risk of inter-species transfer due to mobile genetic elements [51]. This is an area of high priority for future genomic surveillance in The Gambia and elsewhere in West Africa to help guide antimicrobial stewardship and AMR surveillance.

The high maternal MDR-GNB carriage prevalence (76%) is consistent with other African studies [24] and contrasts with lower prevalence observed in Europe (France, 12.8%;) [24] and the Middle East (Lebanon, 19.1%). As maternal samples were obtained within 72h of NNU admission following delivery at different health facilities, we cannot speculate on the source of maternal carriage which may reflect widespread community prevalence or health facility related acquisition during labour. Dissemination of ESBL-GNB in African communities is widespread, with outpatient cross-sectional studies indicating 32.6% carrier prevalence for children in Guinea-Bissau [48], 21.1% prevalence for children in Madagascar [52] and 63.3% carriage prevalence in adults and children in Egypt [53]. Maternal acquisition of GNB temporally related to hospital admission has also been described with rectovaginal carriage prevalence increasing from 18.8% pre-delivery to 41.5% at time of postnatal ward discharge in Sri Lanka [54].

A key finding is that our neonatal cohort carried genetically different MDR-GNB isolates on their skin and intestine compared to paired maternal rectovaginal carriage. We identified only one newborn-mother pair with identical *E. coli* and *K. pneumoniae* strains, despite high maternal and newborn carriage prevalence, suggesting that mothers do not play a prominent role in newborn MDR-GNB acquisition during the perinatal and early post-natal period at this site. This contrasts with evidence from HIC and MIC indicating that maternal MDR-GNB carriage is a risk factor for neonatal acquisition, with estimated 19% (pooled from 5 studies) of neonatal colonisation associated with transmission from colonised mothers [24]. However, there is a lack of robust genomic research examining mother to newborn MDR-GNB transmission in LIC settings, especially Africa [24] and extrapolation of findings from other settings should be avoided, as neonatal MDR-GNB transmission is complex and influenced by multiple context-specific health system factors [11]. A cross-sectional study of Gambian newborns with clinical early onset sepsis reported low prevalence of vertical transmission from maternal genital tract colonisation with only 14% transmission risk for *S aureus* and no genotypically related GNB isolates from mother-newborn pairs [25]. A cohort study in South Africa reported similarly low instances (1.1%) of clonal relatedness between maternal and neonatal derived ESBL-producing *Enterobacter cloacae* [55] and a Sri Lankan paired cohort study reported 0.6% maternal transfer rate for ESBL-producing Enterobacterales [54]. A detailed genomic transmission study conducted in a similar low resource NNU in Madagascar also reported no involvement of family members, including mothers, in transmission [56].

We identified no clonal dissemination of *E. coli* and *K. pneumoniae*, suggesting multiple sources. Environmental contamination of NNUs is well recognised [20] with MDR-GNB able to survive prolonged periods on hands, [20] medical products such as gastric feeding tubes, [57] suction machines [58] water supplies and sinks and on inanimate surfaces [18,59]. Endemic and epidemic MDR-GNB outbreaks occurred at our study site in the year preceding this study, with *Burkholderia cepacia* and ESBL-*K. pneumoniae* isolated from IV fluid preparations and antibiotic vials with genotypic linkage to invasive isolates [22]. As environmental samples were not collected during our study we cannot comment on exact sources of environmental acquisition. However, the absence of evidence for mother to newborn transmission, extensive carriage of MDR-*K. pneumoniae* at 7 days and heterogeneous diversity of strains identified is highly suggestive of multiple environmental sources. This should be confirmed by future research, ideally with linked environmental surveillance from the range of sites at which mother and newborn are managed during labour and the early postnatal period, including place of delivery, referral site and tertiary NNUs.

Limitations of this study include sequencing of single bacterial colonies, which may not have captured the extensive within-host diversity of intestinally carried GNB [60]. Samples were collected over a short period during the dry season and MDR-GNB carriage may differ with seasonality, as shown by other Gambian studies of bacterial infection and carriage [61]. Samples were limited from 14 days onwards due to high mortality and we are unable to comment on the persistence or resolution of MDR-GNB carriage beyond 7 days nor timing of acquisition of maternal carriage.

Further research is required to confirm our findings, with larger sample sizes and linkage to clinical outcomes, including elucidation of the exact timing of acquisition, risk factors for MDR-GNB carriage and association with invasive infections. Our findings suggest that environmental sources play an important role in neonatal MDR-GNB transmission, warranting further targeted study to delineate and identify reservoirs at each point along the newborns’ journey from labour ward to NNU. Exploration of maternal acquisition of MDR-GNB carriage within both community and hospital settings is also needed to identify interventions to interrupt the circulation of these important neonatal pathogens.

## Conclusion

Gambian hospitalised small vulnerable neonates have high carriage prevalence of MDR-and ESBL-GNB with acquisition between birth and 7d of admission. Despite high maternal MDR-GNB carriage prevalence we identified only limited evidence supporting mother to neonate transmission. Heterogeneous diversity of *E. coli* and *K. pneumoniae* strains and extensive AMR gene presence indicates multiple environmental sources from delivery site to neonatal unit. More comprehensive genomic studies of neonatal and maternal MDR-GNB transmission are required to fully understand acquisition pathways in a variety of low resource settings, to inform development of targeted infection prevention control interventions for the most vulnerable newborns.

## Declarations

### Availability of data

Sequence data has been uploaded to the Sequence Read Archive accession number PRJNA73082.

### Competing interests & transparency declaration

No author declared a conflict of interest about the submitted work. BK reports grants from: the MRC UK Research & Innovation (UKRI), United Kingdom; Wellcome Trust, United Kingdom; and Bill and Melinda Gates Foundation (BMGF), United States for a variety of projects related to vaccines and maternal and newborn health. BK attended the Gates Global Challenge Meeting in 2022, supported by BMGF, and is also on the Data Safety Monitoring Board for a COVID producing vaccine company. The other authors declare they have no competing interests.

### Authors contributions

HB and TdS conceptualised the study and obtained funding with input from JEL and BK. Data and sample collection were conducted by BFKK and RB with oversight from HB and JEL. NK and SD performed microbiological processing. MK conducted all DNA extraction and sequencing procedures with input from AK, TdS and AKS. SYB performed all bioinformatic analyses, including generation of phylogenetic trees and MLST analysis. SYB and HB drafted the manuscript and generated the figures with input from MK, SD and TdS. All authors provided input to the overall direction and content of the paper and have seen and approved the final version. SYB and MK contributed equally to this work.

## Data Availability

All genomic data produced are available online at the Sequence Read Archive (accession number PRJNA73082). Clinical meta-data produced in the present study are available upon reasonable request to the authors.

## Acknowledgements

The authors acknowledge the following persons at MRC Unit The Gambia at LSHTM: Yusupha Njie, Binta Saidy and Bai Lamin Dondeh (Data Management); Alpha Jallow, Njilan Johnson, Marie-Rose Thorpe, Elizabeth Batichilly (Research Support). We thank Buntung Ceesay, Mamadou Jallow and Dawda Cham for laboratory contributions and support in addition to Demba Sanneh and Mathurin Diatta (Biobank). In addition, we appreciate the Gambian Government Ministry of Health and Medical Advisory Board at Edward Francis Small Teaching Hospital, for facilitating the field work. Finally, we would like to thank the newborns and their mothers for generously taking part in this study.

