## Supplemental Table 1 for "EARLY ACQUISITION AND CARRIAGE OF GENETICALLY DIVERSE MULTI-DRUG RESISTANT GRAM-NEGATIVE BACILLI IN HOSPITALISED SMALL VULNERABLE NEWBORNS IN THE GAMBIA"

| **Gram-negative bacilli species** | ***Escherichia coli*** | ***Klebsiella pneumoniae*** | ***Enterobacter cloacae*** | ***Acinetobacter baumannii*** | ***Citrobacter freundii*** | ***Salmonella enterica*** | ***Cronobacter*** | ***Pseudomonas aeruginosa*** | ***Pseudomonas putida*** | **Unknown species** | **All gram-negative bacilli** |
| --- | --- | --- | --- | --- | --- | --- | --- | --- | --- | --- | --- |
| **Number of isolates** | **45** | **37** | **7** | **11** | **3** | **1** | **3** | **2** | **1** | **2** | **112** |
| **Beta-lactam (*bla)* genes (N^o^)** | AmpH: 45  PBP: 45  AmpC2: 45  AmpC1: 35  TEM-105: 24  CTX-M-15: 4  CTX-M-27: 2  TEM-1D: 1 | *bla*-AmpH: 36  *bla*-PBP: 35  (bla)CTX-M-15: 20  (bla)TEM-105: 18  (bla)OXA-1: 12  (bla)SHV-187: 12  (bla)SHV-65: 4  (bla)SCO-1: 4  (bla)SHV-75: 3  (bla)SHV-13: 3  (bla)OKP-B-2: 3  (bla)SHV-106: 2  (bla)SHV-27: 2  (bla)SHV-61: 1  (bla)SHV-157: 1  (bla)SHV-1: 1  (bla)OKP-B-3: 1  (bla)OKP-B-19: 1  (bla)LEN-24: 1 | (bla)AmpH: 7  (bla)PBP: 7  (bla)CTX-M-15: 4  (bla)TEM-105: 4  (bla)ACT-16: 3  (bla)AZECL-25: 3  (bla)OXA-1: 3  (bla)ACT-7: 1 | (bla)Mbl: 10  (bla)BlaA1: 9  (bla)BlaA2: 6  (bla)beta-lactamase_class-C: 4  (bla)ADC-68: 3  (bla)ADC-77: 3  (bla)CARB-5: 2  (bla)OXA-91: 2  (bla)OXA-64: 2  (bla)OXA-117: 1  (bla)OXA-120: 1  (bla)OXA-180: 1 | (bla)AmpH: 3  (bla)PBP: 3  (bla)TEM-105: 1  (bla)AmpC1: 1  (bla)AmpC2: 1  (bla)CMY-70: 1  (bla)CMY-79: 1  (bla)VEB-1: 1 | (bla)AmpH: 1  (bla)PBP: 1  (bla)TEM-105: 1  (bla)AmpC1: 1  (bla)AmpC2: 1 | (bla)AmpH: 3  (bla)PBP: 3 | (bla)OXA-50: 2 | - | (bla)CTX-M-15: 1  (bla)DHA-1: 1  (bla)OXA-1: 1 |  |
| Total N^o^ (bla) genes | 201 | 160 | 32 | 44 | 12 | 5 | 6 | 2 | - | 3 | **465** |
| **Aminoglycoside (AGly) genes (N^o^)** | StrA: 31  StrB: 31  AadA5: 8  AadA1-pm: 4  Sat-2A: 2  Aac3-IId: 1  AadA2: 1 | (AGLy)StrB: 21  (AGLy)StrA: 20  (AGly)Aac3-IIa: 18  (AGly)AadA1-pm: 5  (AGly)AadA16: 4  AGly)AadA2: 2  (AGly)AadA3: 1  (AGly)Aph3-Ia: 1  (AGly)Aac3-IId: 1 | (AGLy)StrA: 4  (AGLy)StrB: 4  (AGly)AadA1-pm: 4  (AGly)Aac3-IIa: 3 | (AGly)Aph3-Ia: 2  (AGly)AadA1-pm: 2  (AGly)AadB: 2  (AGly)Sat-2A: 2  (AGLy)StrA: 1  (AGLy)StrB: 1 | AGLy-StrA: 1  AGLy-StrB: 1  AGly-AadA24: 1  AGly-AadA5: 1  AGly-AadB: 1 | AGLy-StrA: 1  AGLy-StrB: 1  (AGly)Aac3-IId: 1  (AGly)AadA5: 1 | - | AGly-Aph3-IIb: 2 | AGly-AadA5: 1  MLS-MphA: 1 | (AGly)Aph3-Ib: 1  (AGly)Aph6-Id: 1  (AGly)Aac3-IId: 1  (AGly)Aac6-Ib: 1 | **-** |
| Total N^o^ (Agly)genes | 78 | 73 | 15 | 10 | 5 | 4 | - | 2 | 2 | 4 | **193** |
| **Trimethoprim (Tmt) genes (N^o^)** | Dhfr7: 10  DfrA17: 8  DfrA14: 4  DfrA1: 4  DfrA8: 3  DfrA5: 2  DfrA12: 1 | (Tmt)DfrA14: 14  (Tmt)Dfr15b: 5  (Tmt)DfrA27: 4  (Tmt)DfrA12: 2  (Tmt)Dhfr7: 1  (Tmt)Dfr16: 1  (Tmt)Dfr22:1 | (Tmt)DfrA14: 2  (Tmt)Dfr15b: 1 | (Tmt)DfrA1: 2 | (Tmt)DfrA14: 1  (Tmt)Dfr15b: 1  (Tmt)DfrA17: 1 | (Tmt)DfrA17: 1 | - | - | (Tmt)DfrA17: 1 | (Tmt)DfrA1: 1 | **-** |
| Total N^o^ (Tmt) genes | 32 | 28 | 3 | 2 | 3 | 1 | - | - | 1 | 1 | **71** |
| **Chloramphenicol**  **(Phe) genes (N^o^)** | CatA1: 7 | (Phe)CatA1: 2  (Phe)CatA2: 6  PheCatB4: 15 | (Phe)CatA1:3  (Phe)CatB4: 3 | (Phe)FloR: 1  PheCmlB1: 1 | (Phe)CatA1: 1  (Phe)CmlA5: 1 | - | - | (Phe)CatB7: 2 | - | (Phe)CatA2: 1 | **-** |
| Total N^o^ (Phe) genes | 7 | 23 | 6 | 2 | 2 | - | - | 2 | - | 1 | **43** |
| **Quinolone (Flq) genes (N^o^)** | Qnr-S1: 6 | Flq*OqxA*: 33  Flq*OqxBgb*: 33  (Flq)QnrB1: 12  (Flq)Qnr-S1: 4  (Flq)QnrB6: 4 | (Flq)*OqxA*: 6  (Flq)*OqxBgb*: 6  (Flq)QnrB1: 3 | - | (Flq)QnrB34: 1  (Flq)QnrVC4: 1 | - | - | - | - | **-** | **-** |
| Total N^o^ (Flq) genes | 6 | 86 | 15 | - | 2 | - | - | - | - | **-** | **109** |
| **Tetracycline (Tet) genes (N^o^)** | Tet-34: 31  TetR: 20  TetA: 20  TetB: 3  TetD: 1 | TetTetR: 15  TetTetA: 15  TetTetD: 6 | TetTetR: 3  TetTetA: 3 | (Tet)TetB: 2 | TetTetR: 2  TetTetA: 2  TetTet-34: 1 | TetTetR: 1  TetTetA: 1 | - | - | - | (Tet)tet39: 1  (Tet)tetB: 1 | **-** |
| Total N^o^ (Tet) genes | 75 | 36 | 6 | 2 | 5 | 2 | - | - | - | 2 | **128** |
| **Sulfonamide (Sul) genes (N^o^)** | SulII: 29  SulI: 21 | (Sul)*sulII:* 20  (Sul)*sulI:* 12 | (Sul)*sulII: 4*  (Sul)*sulI: 1* | (Sul)SulII: 3 | (Sul)SulI: 2  (Sul)SulII: 1 | (Sul)SulI: 1  (Sul)SulII: 1 | - | - | (Sul)SulI: 1 | (Sul)SulI: 1  (Sul)SulII: 1 | **-** |
| Total N^o^ (Sul) genes | 50 | 32 | 5 | 3 | 3 | 2 | - | - | 1 | 2 | **98** |
| **Macrolides (MLS) genes (N^o^)** | MphA: 7 | (MLS)MphA: 2 | - | (MLS)MphE: 1  (MLS)MsrE: 1 | (MLS)MphA: 1 | - | - | - | - | **-** | **-** |
| Total N^o^ (MLS) genes | 7 | 2 | - | 2 | 1 | - | - |  | - | **-** | **12** |
| **Fosfomycin (Fcyn) genes (N^o^)** | - | (Fcyn)FosA: 2 | (Fcyn)FosA2: 6 | - | - | - | - | - | - | **-** | **-** |
| Total N^o^ (Fcyn) genes | - | 2 | 6 | - | - | - | - | - | - | **-** | **8** |
| **Rifampicin**  **(Rif) genes (N^o^)** | - | Arr3: 4 | - | - | - | - | - | - | - | **-** | **-** |
| Total N^o^ (Rif) genes | - | 4 | - | - | - | - | - | - | - | **-** | **4** |
| **TOTAL AMR GENES** | **456** | **446** | **88** | **75** | **33** | **14** | **6** | **6** | **4** | **13** | **1141** |

**Supplementary table 1. Antimicrobial resistance genes identified from neonatal and maternal GNB carriage isolates**
